# Laparoscopic vs open gastrectomy- An updated metanalysis of randomized control trials for short term outcomes

**DOI:** 10.1101/2020.04.12.20062562

**Authors:** Bhavin Vasavada, Hardik Patel

**Author notes:** Financial disclosures: none.

## Abstract

**Introduction:** Aim of this metanalysis was to compare short term outcomes of laparoscopic and open gastrectomy for gastric cancer.

**Material and methods:** EMBASE, MEDLINE, PubMed and the Cochrane Database were searched for randomized control trials comparing outcomes in patients undergoing laparoscopic gastrectomies with those patients undergoing open gastrectomies. The primary outcome was 30 days morbidity and mortality. Secondary outcomes studied included length of stay, blood loss, d2gastrectomies, lymph node retrieval, operative time, distal gastrectomy, wound complications and intraabdominal complications Systemic review and Metanalysis were done according to MOOSE and PRISMA guidelines.

**Results:** Morbidity was significantly low in laparoscopic group (P=0.003).There was no significant difference between mortality between the two groups. (P=0.75). There fewer wound complications in laparoscopic group, no difference intra-abdominal complications in both the groups. Blood loss was significantly lesser in laparoscopic group.(p <0.001). Hospital stay was similar in laparoscopic group. (P=0.30). Operative time was significantly higher in laparoscopic group. (P< 0.001). Laparoscopic group patients had less number of lymph node retrieval compared to laparoscopic group.(p = 0.002). Laparoscopic group also contained similar advanced staged gastric cancer than open gastrectomies. (p= 0.64)

**Conclusions:** Laparoscopic gastrectomies were associated with better short term outcomes.

## Introduction

With advancement of technology and skills laparoscopic gastrectomy is increasingly being performed. However, there is still some debate over short term outcome, oncologic safety of resections and long-term survivals in comparison to standard open gastrectomies.[1]. Initially laparoscopic gastrectomy was reserved for early and distal laparoscopies but these days more and more surgeons are performing laparoscopic gastrectomies via open approach also. [2].

### AIMS OF STUDY

Aims of this metanalysis to do analysis of recent randomized control trials regarding short-term outcomes.

In short term outcomes aim was to study morbidity and in hospital mortalities as wells as hospital stay, blood loss, operative times as well as to study oncological parameters like D2gastrectomies, number of lymph node retrieval D2gastrectomies, number, resection rates for advanced gastric cancers.

## Material and methods

EMBASE, MEDLINE, PubMed and the Cochrane Database were searched for randomized control trials comparing outcomes in patients undergoing laparoscopic gastrectomies with those patients undergoing open gastrectomies and studies comparing long-term survival outcomes. Two independent authors extracted the data (B.V and H.P). Discussions and mutual understanding resolved any disagreements. Systemic review and Metanalysis was done according to MOOSE and PRISMA guidelines. [14,15]. Types of studies included in metanalysis is described in table 1.

**Table 1.** charecteristics of studies for short term out comes

| Study | Type of study | Number of patients<br>laproscopic<br>gastrectomy | Number of patients<br>in open group |
| --- | --- | --- | --- |
| KLASS02 | RCT | 513 | 498 |
| Cristiano<br>2005 | RCT | 30 | 29 |
| HU<br>2016 | RCT | 519 | 520 |
| LEE 2004 | RCT | 24 | 23 |
| HIYASHI 2005 | RCT | 14 | 14 |
| KITANO 2002 | RCT | 14 | 14 |
| KIM<br>2016 | RCT | 686 | 698 |
| Cia<br>2011 | RCT | 49 | 47 |
| Takiguchi<br>2013 | RCT | 20 | 20 |
| Cui<br>2015 | RCT | 128 | 142 |
| katai2017 | RCT | 455 | 157 |

### Statistical analysis

The meta-analysis was conducted using Open meta-analysis software. Heterogeneity was measured using Q tests and I^2^, and p < 0.10 was determined as significant (8). If 2 there was no or low heterogeneity (I^2^ < 25%), then the fixed-effects model was used. Otherwise, the random-effects model was used. The Odds ratio (OR) was calculated for dichotomous data, and weighted mean differences (WMD) were used for continuous variables. Both differences were presented with 95% CI. For continuous variables, if data were presented with medians and ranges, then we calculated the means and Standard deviations according to Hozo et al. (16). If the study presented the median and inter-quartile range, the median was treated as the mean, and the interquartile ranges were calculated using 1.35 SDs, as described in the Cochrane handbook.

### Inclusion criteria for studies

- Randomized control trials for short term outcomes.
- Studies comparing laparoscopic and open gastrectomies.
- Full text articles.

### Exclusion criteria for studies

- Nonrandomized control trials for short term outcomes.
- Studies with single groups or studies in which groups were not comparable.
- Studies where full texts were not available
- Duplicate studies.

### Assessment of Bias

Characteristics of the studies are described in table 1. [3-13,19-26]. Randomized trials were assessed based on the Cochrane Handbook. [18] (Figure 2). We evaluated publication bias by funnel plots for each parameter.

## RESULTS

Selection process of studies for short term and long-term outcomes for this meta-analysis is described in Figure 1.

**Figure 1:**
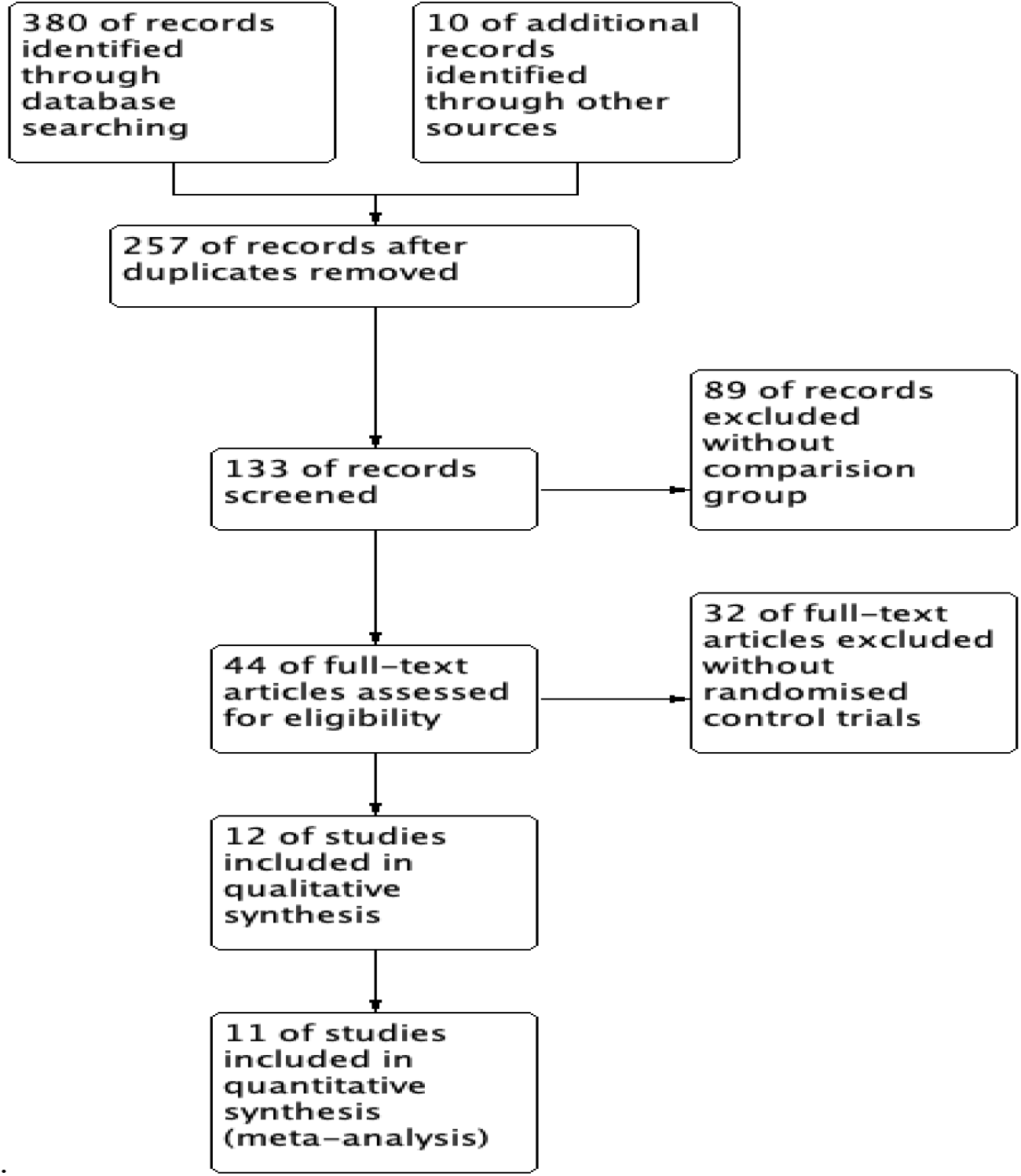
Search strategy according to PRISMA guidelines for short term outcomes

**Figure 2:**
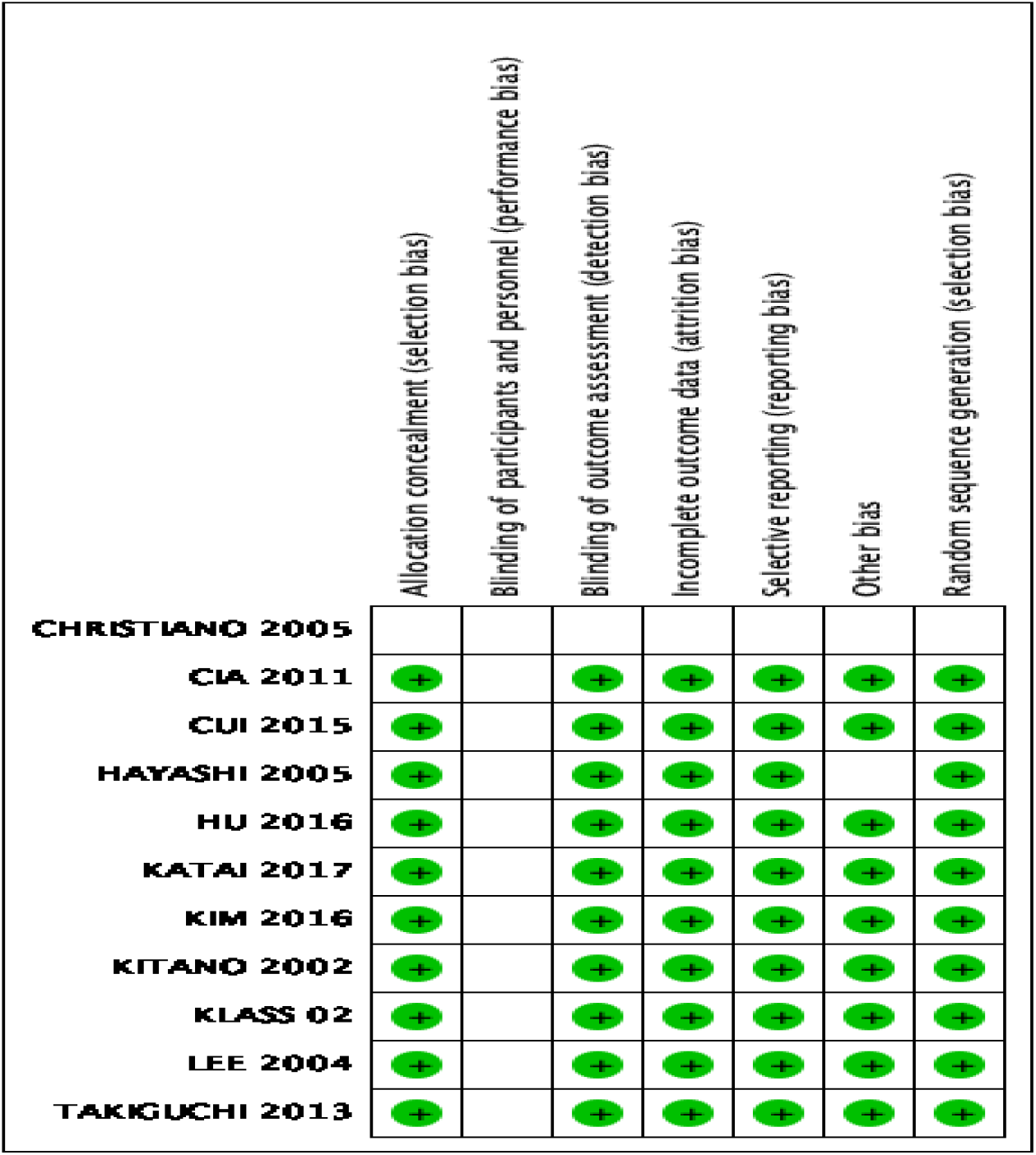
Risk of bias summary of RCT. + denotes low risk of bias, – denotes high risk of bias.

For short-term outcomes 11 RCTS consisting of 4614 patients were included in study. Total 2452 patients were there in laparoscopic gastrectomy group while 2162 patients were included in open gastrectomy group. Morbidity is defined as any deviation from normal perioperative course.

Postoperative morbidity was significantly low in laparoscopic group. (P= 0.03). There was no significant difference between mortality between the two groups. (P=0.75). [figure 3]

**Figure 3.**
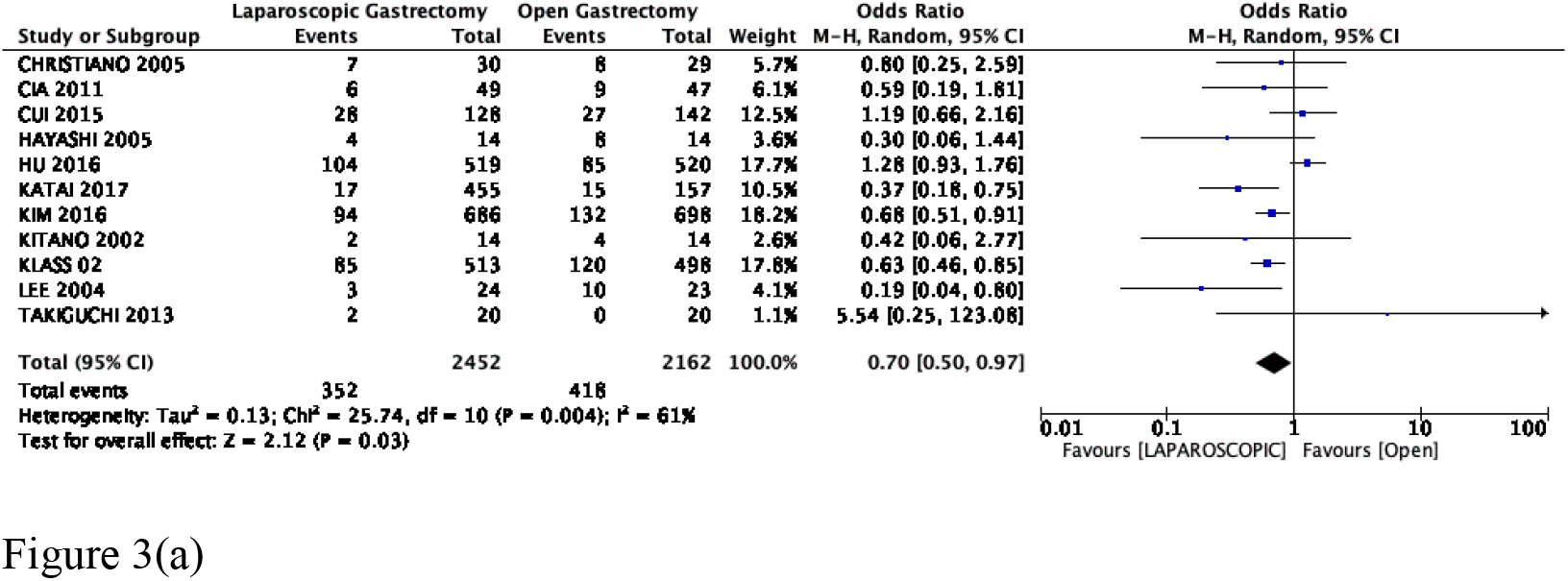
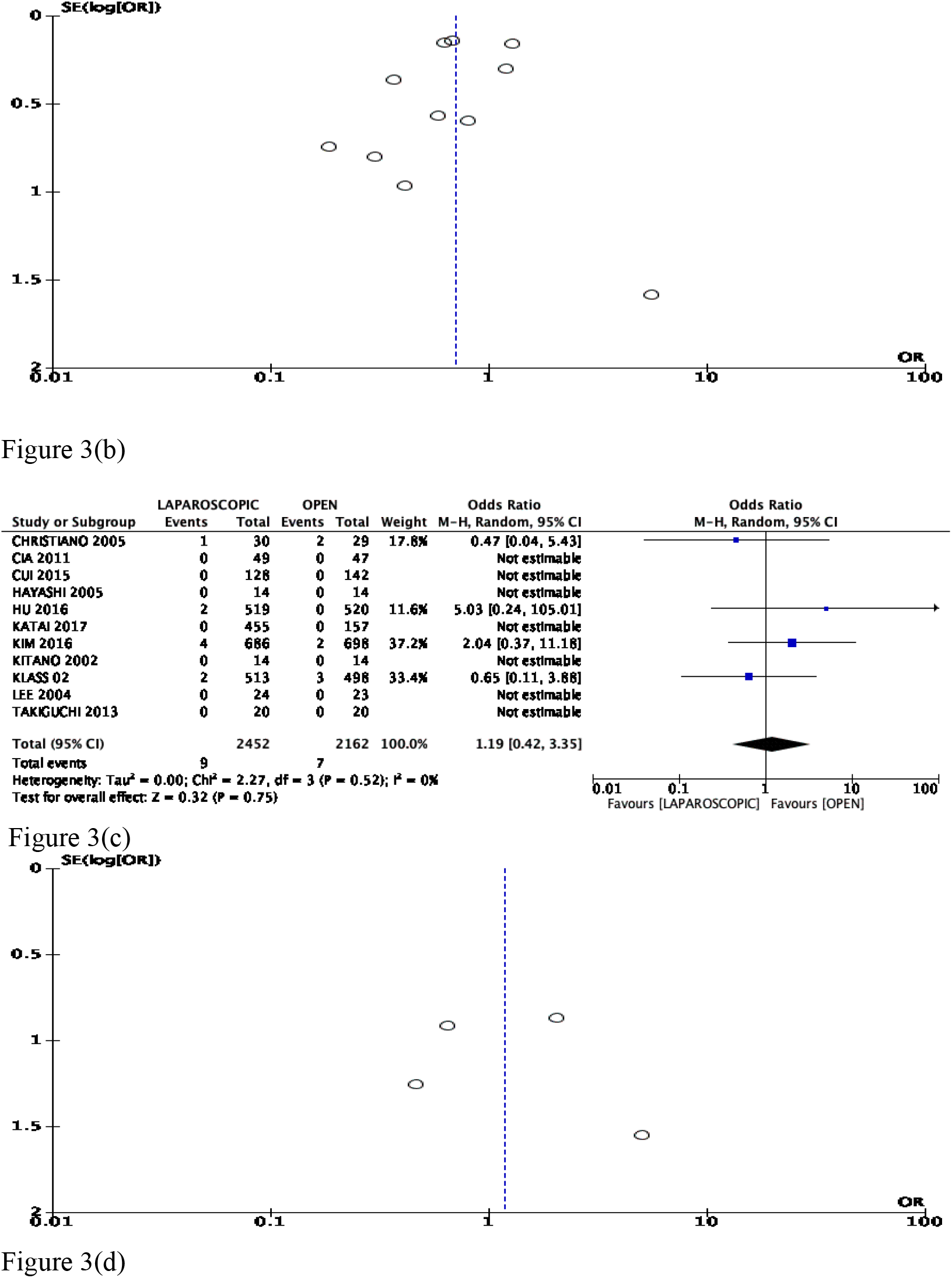
(a)Comparision between morbidity laproscopic and open gastrectomy. (b) Funnel point of morbidity (c) Forest plot of mortality between laparoscopic and open gastrectomy (d) Funnel plot of mortality.

Wound complication was significantly less in laparoscopic group (p=0.009), other intra-abdominal complications were in both the groups. (P=0.18). [figure 4]

**Figure 4.**
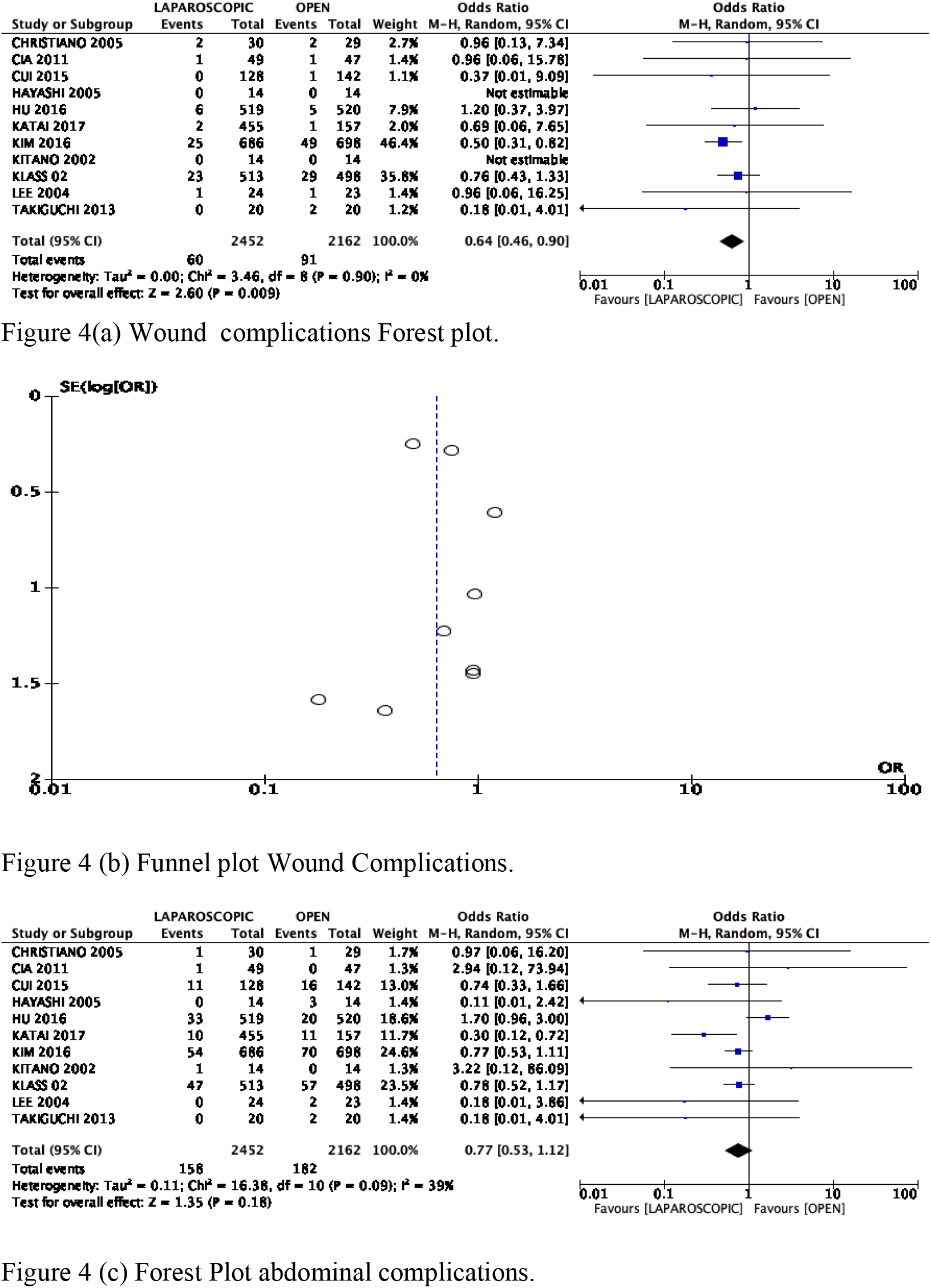
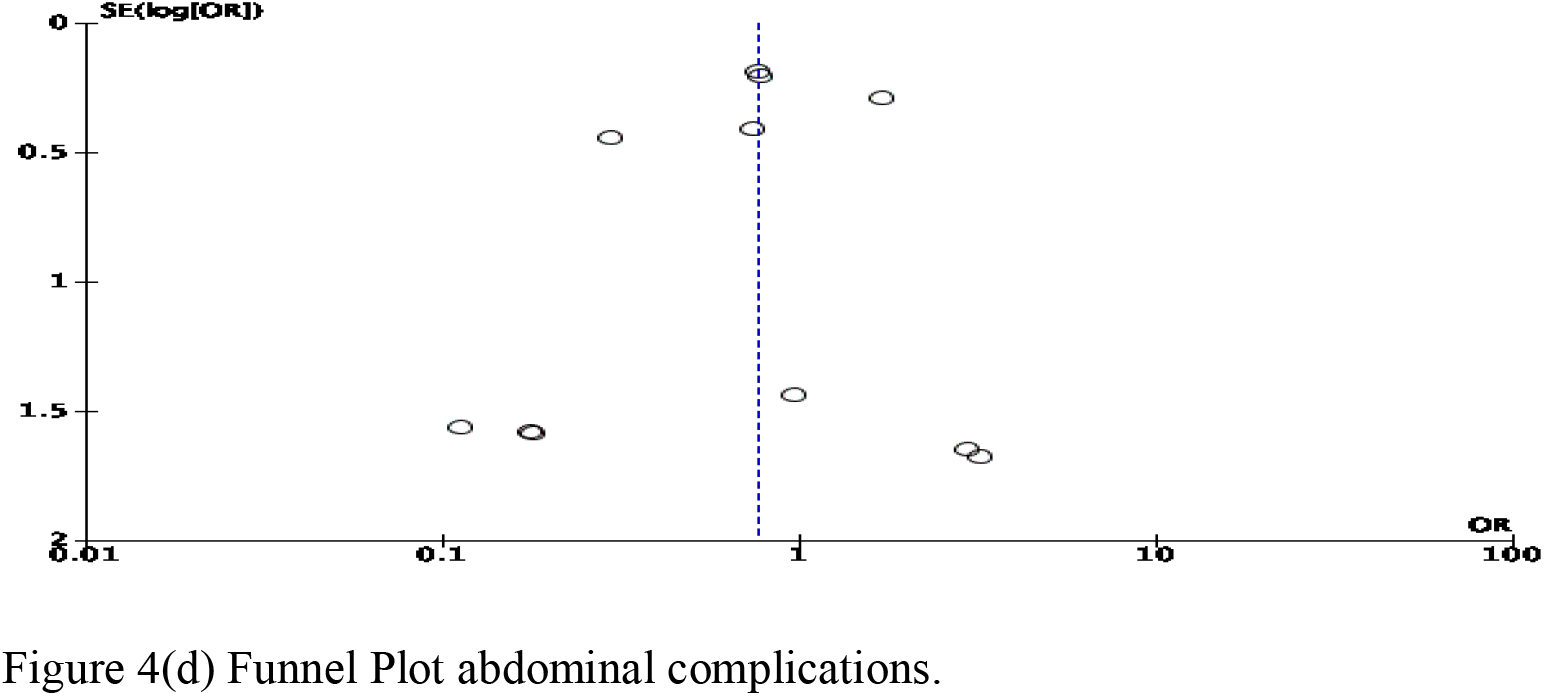
wound complications was significantly less but no difference in intraabdominal complications.

Blood loss in ml was significantly lesser in laparoscopic group.(p <0.001). Hospital stay in days was not significantly different in laparoscopic group. (P=0.30). Operative time in minutes was significantly higher in laparoscopic group. (P< 0.001). [figure 5] Laparoscopic group patients had similar d2 gastrectomies (p=0.26) and less number of lymph nodes retrieved compared to laparoscopic group.(p=0.002), Laparoscopic group also undergone similar advanced staged gastric cancer (T2 and higher) than open gastrectomies. (p=0.640). [figure 6]

**Figure 5.**
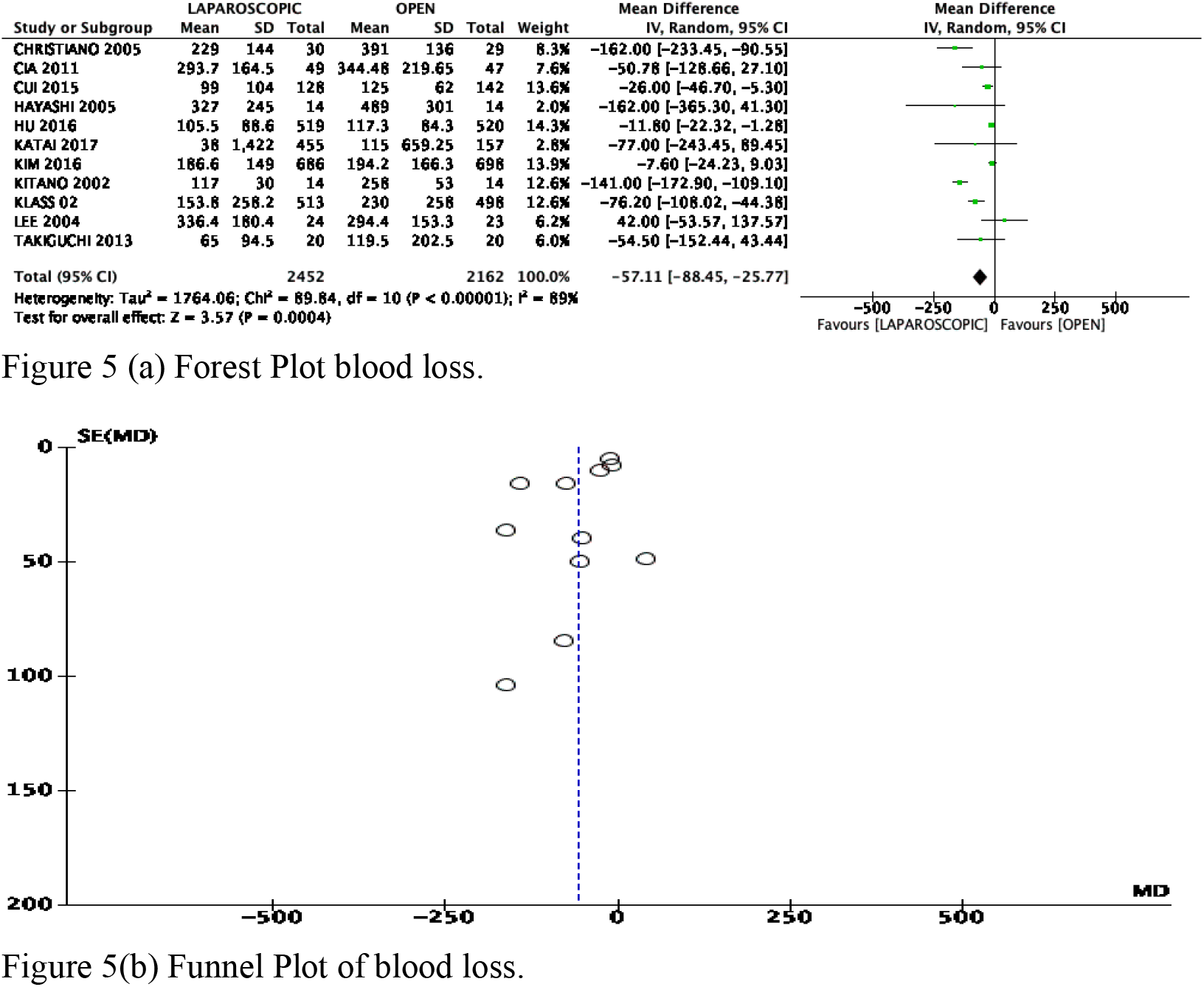
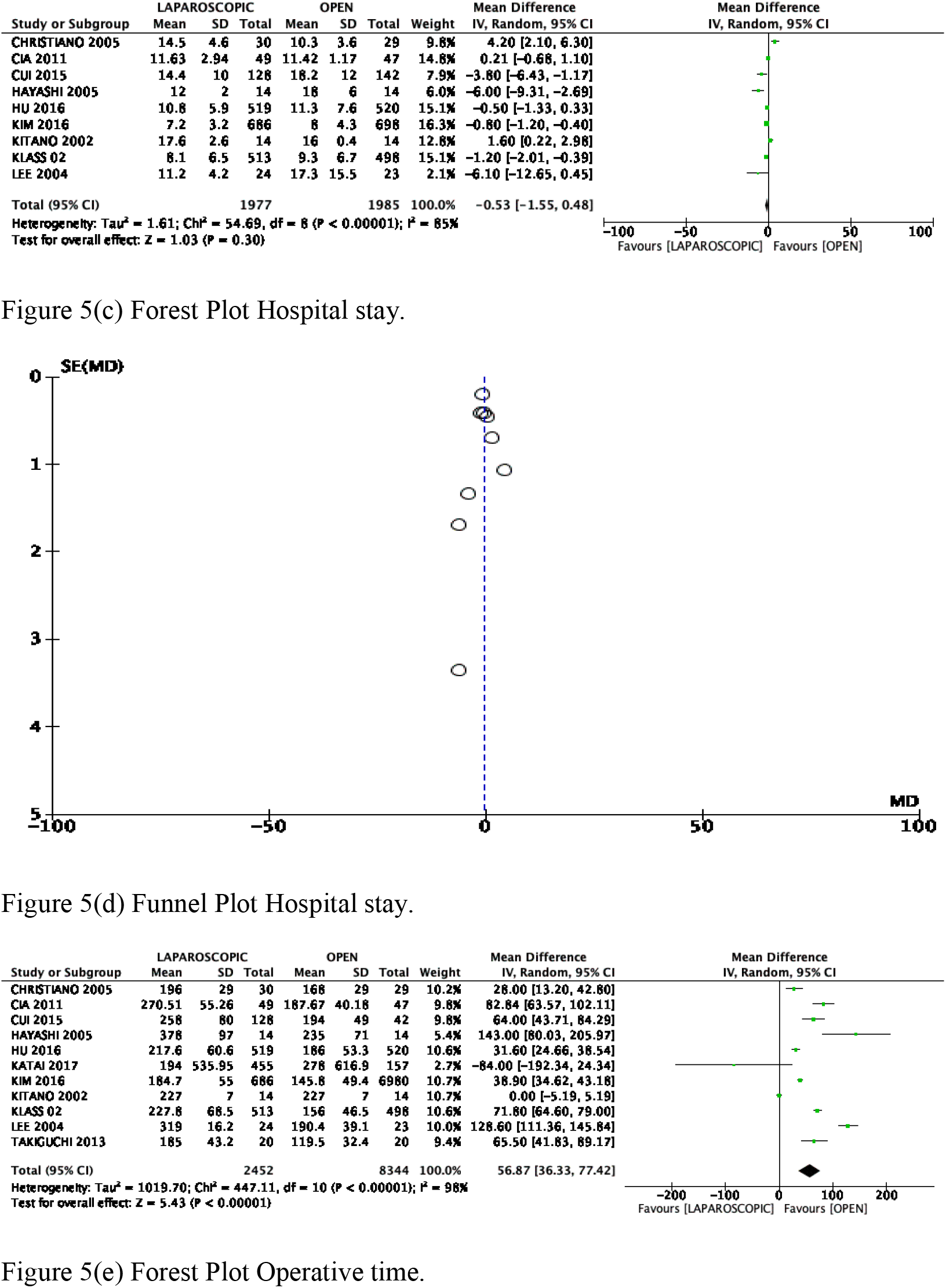
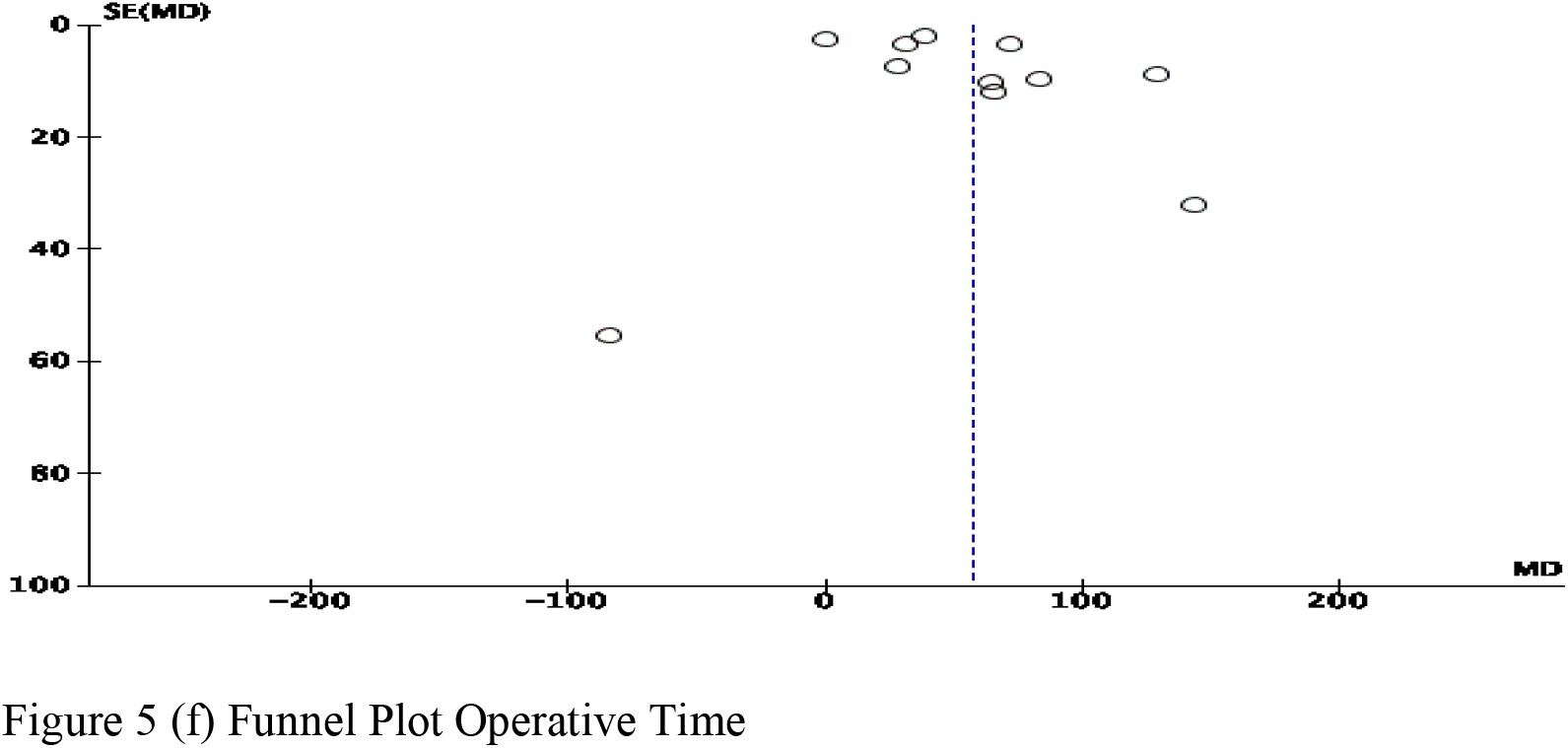
Comparisons of blood loss, hospital stay and operative time between laproscopic and open gastrectomy.

**Figure 6.**
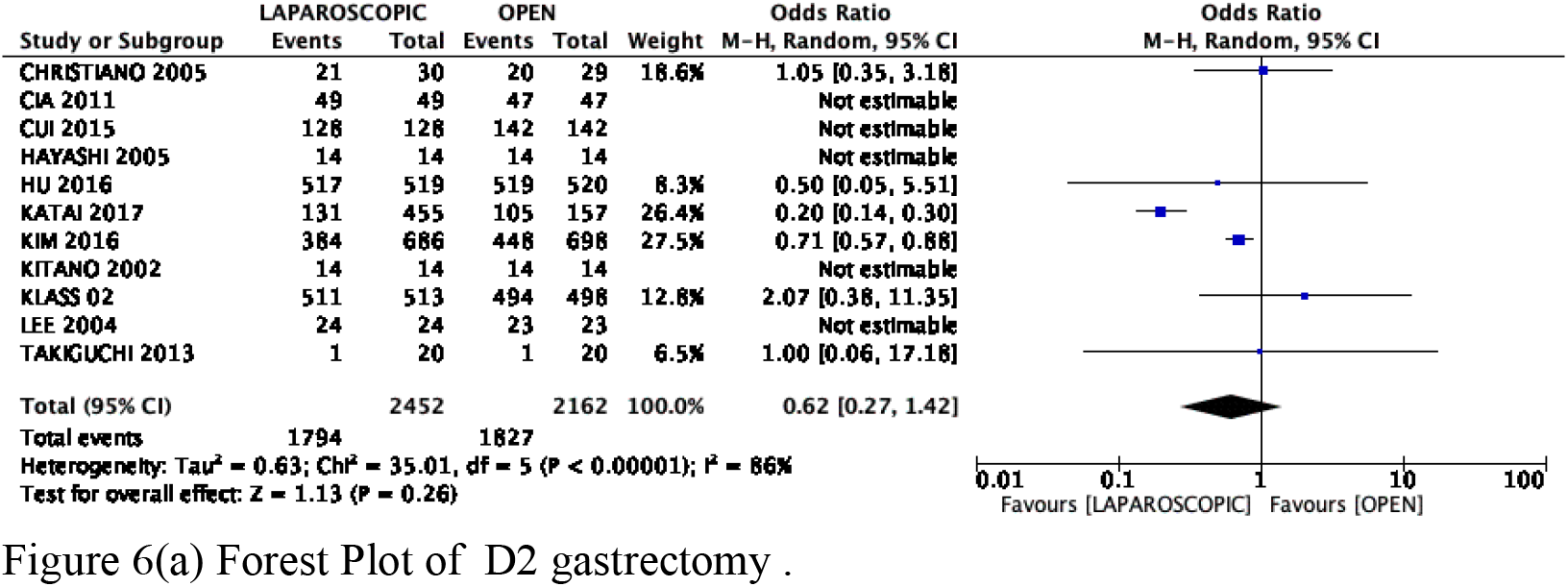
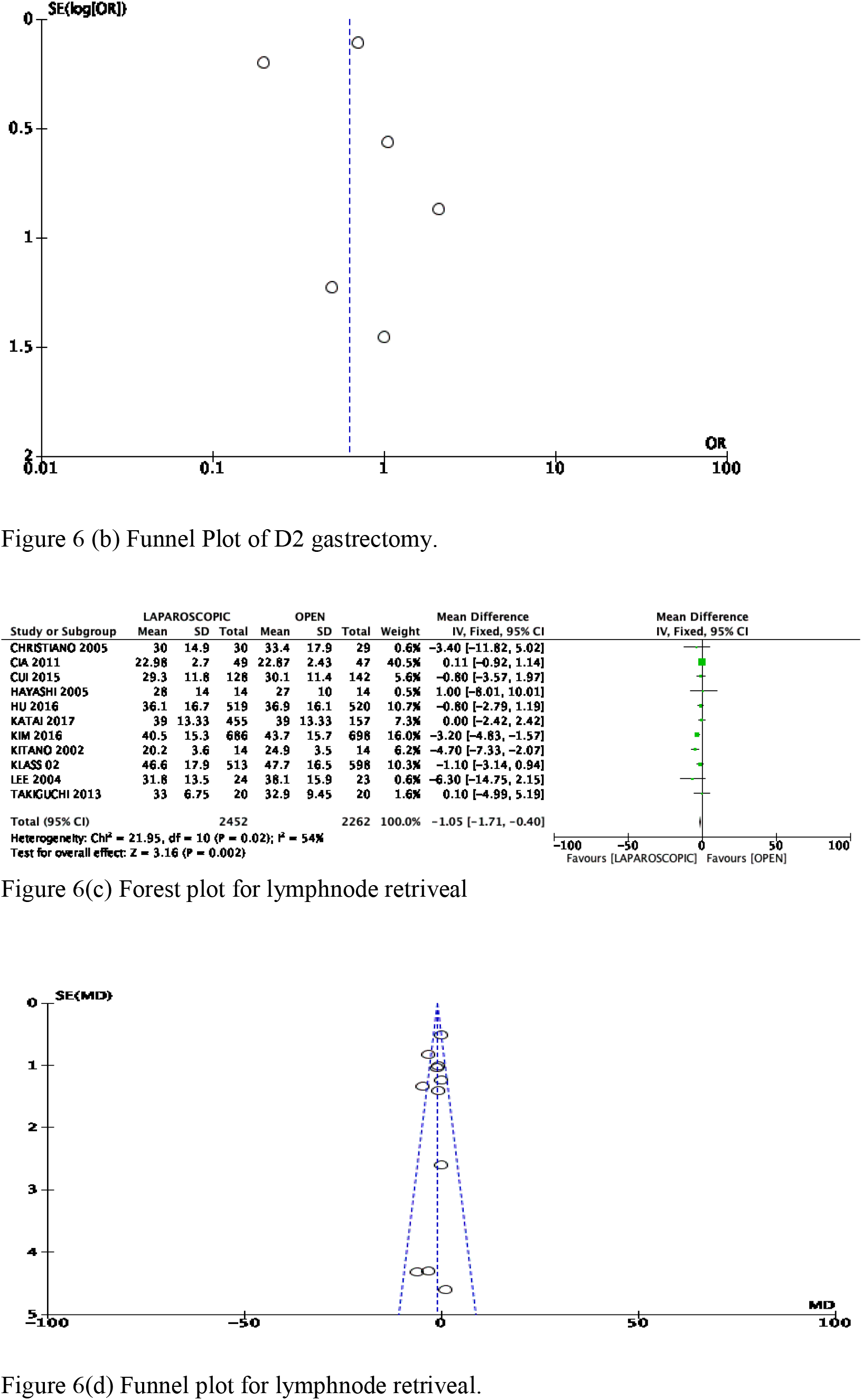
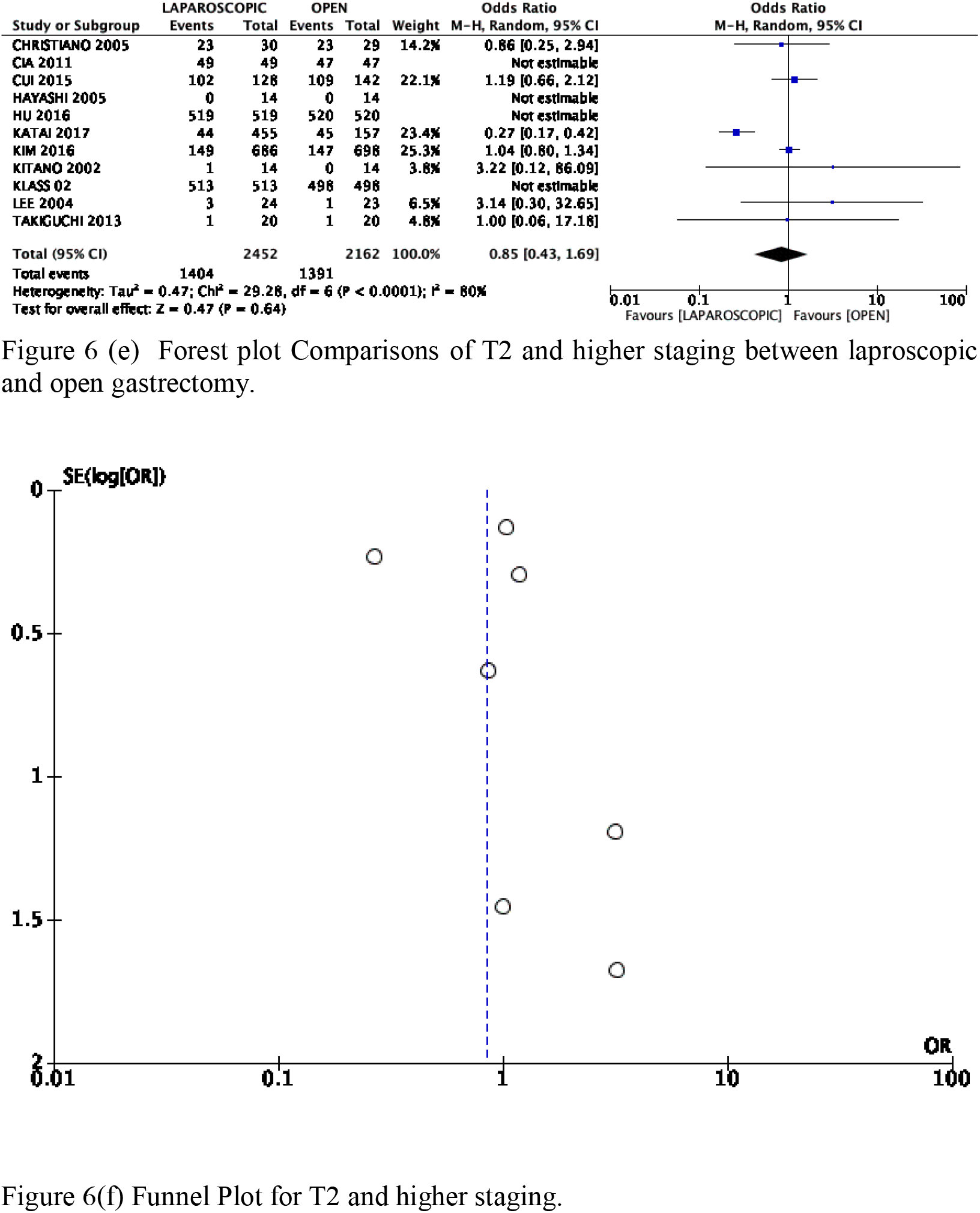
comparision of completion of d2 gastrectomy,number of lymphnode retrived and T2 and high staging between laproscopic and open gastrectomy

## DISCUSSION

Laparoscopic surgeries have shown similar results to open surgery with improved perioperative outcomes in many malignant diseases. [26-30]. With time laparoscopic approaches have gained more popularity for gastric cancers. [31-32]

Aim of our study was to perform updated meta-analysis of recent randomized control trials for short term outcomes comparing laparoscopic vs open gastrectomies. For long term out comes there are not enough randomized control trials for long term survival outcomes. So we performed meta analysis of cohort studies for long term survivals.

90 days morbidity was significantly less in laparoscopic group however on subgroup analysis there was no difference in Local complications (leak, fistula, collection), wound complications (SSI) were significantly lesser in laparoscopic group. Our findings do suggest that benefit of laparoscopic surgeries in short term morbidities..

There was no difference in 90 days mortality between the two groups. Operative time was significantly higher in laparoscopic group, intraoperative blood loss was significantly less in laparoscopic group, suggesting that laparoscopic surgery is beneficial in short term outcomes. However there was no significant difference in hospital stay.

There has always been doubt about oncologic safety and adequacy of laparoscopic surgeries. Our meta analysis also gave similar findings, There was significantly lesser number of total number of lymph node retrieval in laparoscopic group. Many recent studies showed oncologic benefit of d2 vs d1 gastrectomies. [33,34]. Long term impact of these findings needs to be evaluated.

There are certain limitation in our analysis. Heterogeneity was significant in some parameters as seen in some forest plot. Some parameters showed publication bias as shown by funnel plots.

In conclusion Laparoscopic gastrectomies were associated with better short term outcomes. However long-term survival rate needs to be evaluated by randomized control trials and further metanalysis.

## Data Availability

meta analysis- data collected from published paper

## Abbreviations

RCT: (randomized control trial)
WMD: Weighted Mean Difference
C.I: Confidence Intervals.
SD: (Standard deviations)

